# Toward early detection of burnout: A systematic review of potential biomarkers

**DOI:** 10.1101/2025.08.05.25332953

**Authors:** Maddalena Balia, Franck Zenasni, Maryne Lepoittevin, Renzo Bianchi, Adrien Julian, Sylvain Bodard, Marie Bringer

**Author notes:** **Corresponding author:** Dr Marie Bringer.

## Abstract

Burnout Syndrome (BOS), a pervasive occupational phenomenon stemming from unmanaged chronic workplace stress, leading to physical, psychological and cognitive impairment, represents a major challenge for preventive medicine. Indeed, the worldwide increasing incidence of BOS and the importance of its early management points to the unmet need for early BOS diagnosis. Whilst biomarkers of chronic stress have been explored with the description of the allostatic load, BOS lacks a consistent physiological signature, which would contribute to an early and comprehensive identification of persons at risk. This systematic review synthesizes current evidence on BOS-related biomarkers, aiming to identify potential physiological correlates. We conducted a comprehensive search of PubMed and EMBASE, yielding 111 studies evaluating 36 biomarkers in adult populations. Our analysis revealed inconsistent associations across most physiological systems, including the hypothalamic-pituitary-adrenal axis (*e.g*., cortisol, DHEA), immune system, cardiovascular parameters. While some biomarkers like HbA1c, blood glucose, or comorbidities like irritable bowel syndrome showed more consistent positive correlations with BOS, the overall findings are largely inconclusive. We conclude that the current biological evidence is insufficient for establishing a definitive BOS biosignature for routine clinical diagnosis. Future research should prioritize a more unified and comprehensive definition of BOS, potentially integrating emerging assessment tools to advance the objective identification and early intervention of burnout.

## Introduction

Burnout syndrome (BOS) t is defined by the World Health Organization (WHO) as a syndrome resulting from chronic, unmanageable workplace stress (1). Its rampant progression worldwide represents an increasingly major health challenge, as a study reports that more than a third of healthcare workers suffer from burnout worldwide (2). BOS is not recognized as a medical disorder in the DSM-5, and is explicitly described in the International Classification of Diseases 11th Revision as an *occupational phenomenon*, not a medical condition. This in-between status likely contributes to the heterogeneity of diagnostic approaches and the ongoing difficulty in identifying consistent biological correlates (3). Even though the definition itself of BOS remains fluctuating, multidimensional, and highly dependent on the analytical framework (clinical, organizational, sociological, etc.) (4,5), the symptoms more often associated with BOS are emotional exhaustion (EE), depersonalisation (DEP) or cynicism, and reduced personal accomplishment (PE) (6). Recently, studies from Schaufeli et al. added cognitive impairment and emotional impairment to the conceptualisation of BOS (7,8). BOS is thought to develop gradually, often preceded by a transitional phase of early BOS defined by chronic and uncompensated stress, leading to a progressive decline in physical, emotional, and cognitive capacities (9). There are currently no established diagnostic criteria for BOS. Symptom assessment remains primarily clinical and often supported by self-reported questionnaires such as the Maslach Burnout Inventory (MBI) (6) or more recently by the Burnout Assessment Tool (BAT) (7). However, the self-reported nature of these tools may limit reproducibility and objectivity (10). Identifying physiological biomarkers could enable a more standardized and clinically applicable approach, supporting early detection and complementing current subjective assessments, which can lead to under or overestimated diagnosis. In order to identify patients on this trajectory and offer them preventive solutions, it is crucial to characterize the biological and psychological signature of BOS. As BOS is assumed to result from chronic workplace stress, biomarkers of allostatic load (AL) are frequently assessed in research on the physiological features of BOS (3). While AL provides a general picture of the physiological and anatomic costs of chronic stress, they may not fully capture the specificity of burnout as a work-related syndrome. A previous systematic review conducted in 2011 evaluated the relationship between a large number of AL and more generally stress-related biomarkers with BOS (3). They couldn’t conclude in any clear association because of a lack of concurrent studies (3). Given the growing demand for objective diagnostic tools in occupational medicine and mental health care, this review aims to ascertain whether specific biomarkers of BOS have been identified. The identification of reliable biomarkers for BOS could be used in complement to self-report questionnaires, in order to enable the development of stratified prevention strategies and personalized therapeutic approaches, adapted to individual physiological profiles and stress resilience.

## Material & methods

### Data source and strategy

Articles research was led on PubMed (http://www.ncbi.nlm.nih.gov/entrez/query.fcgi) and EMBASE (http://www.ovid.com) for the period from inception to June 17, 2025. The search strategy included a combination of predefined keyword terms related to biomarkers and Medical Subject Heading (MeSH) terms for burnout. Studies were eligible for inclusion if they met the following criteria: (1) original studies including observational studies, correlational studies, random and non-random studies in humans, (2) published in English, (3) studies evaluating the association/correlation between biomarkers and burnout, comparing burnout patients to healthy controls or patients with low, moderate and high scores of burnout. We considered only studies in which burnout was adequately described and assessed in either clinical or non clinical adult populations, and we excluded interventional studies. Studies involving pediatric or adolescent populations were excluded. Burnout assessment and stages of burnout had to be determined with validated psychometric questionnaires scores. Reference lists of the retrieved articles were examined for further studies. The complete search strategies can be found in the Supplementary Table 1. This systematic review was registered in PROSPERO (CRD420251075694) and conducted in line with the Preferred Reporting Items for Systematic Reviews and Meta-Analyses (PRISMA) guidelines (Supplementary Table 2) (11).

### Data Extraction

Duplicates were removed, and a first screening based on titles and abstracts were performed independently by two investigators (M.B., M.L.). Any disagreements on article screening were resolved by discussing with another investigator (S.B.). Subsequently, the following information was extracted from full-text using a predefined data-extraction spreadsheet by the same two investigators, comprising the study title, year, biomarker(s) discussed, burnout assessment, level of burnout (early, moderate, severe) assessment, experimental design, sample size, age range, association with biomarkers, considered confounders. Any disagreements on data extraction were resolved by discussing with another investigator (S.B.).Due to the wide variability in study designs, definitions of burnout, and biomarker assessment methods, a formal risk of bias evaluation was not included. Given the descriptive nature of this review and the absence of a meta-analytic component, such an assessment was considered unlikely to provide meaningful additional insights.

### Data Synthesis

Based on the statistical elements of each study, we determined the direction of the association between each biomarker and BOS, which was classified as “positive”, “negative” or “not associated”. We analysed biomarkers that were evaluated in at least two studies. The consistency of association for a given biomarker was then calculated as the proportion of the most frequent direction of association over the total number of reported associations across included studies.

We did not perform any meta-analysis for several reasons. There was no consistency in the self-report questionnaire used for BOS diagnosis -either in the type or the variant used (*e.g.* for MBI, variations on the number of items, dimensions included or target population), the sampling method (*e.g.* for cortisol, salivary, hair or plasmatic, or different time points for cortisol awakening response) or large differences in sample size (Supplementary Table 3).

## Results

1080 articles were identified through our keyword-based search strategy. We screened the abstracts and titles, 223 articles were retained for full-text evaluation. Further investigation and data extraction, 111 studies met all eligibility criteria and were included in the final analysis (Figure 1, Supplementary Table 3). Across the included studies, we analyzed the association of 36 biomarkers with BOS (Table 1). We also reviewed the burnout assessment tools used in the included articles (Table 2). The Maslach Burnout Inventory (MBI) is the most commonly used, as seen in 66.36% of included articles, followed by the Shirom–Melamed Burnout Questionnaire (SMBQ) with 15.89% of articles concerned. In 29 articles, biomarkers association was evaluated in BOS patients and controls, whilst 82 of studies investigated the biomarkers association for low, moderate and high BOS score patients or continuous BOS score evaluation. Amongst the evaluated biomarkers were identified primary and secondary mediators of hypothalamic–pituitary–adrenal axis (HPA) response, immune system, cardiovascular, gastrointestinal, obesity, and other biomarkers. For each biomarker, we determined the frequency, direction and consistency of association with BOS (Table 1).

**Figure 1.**
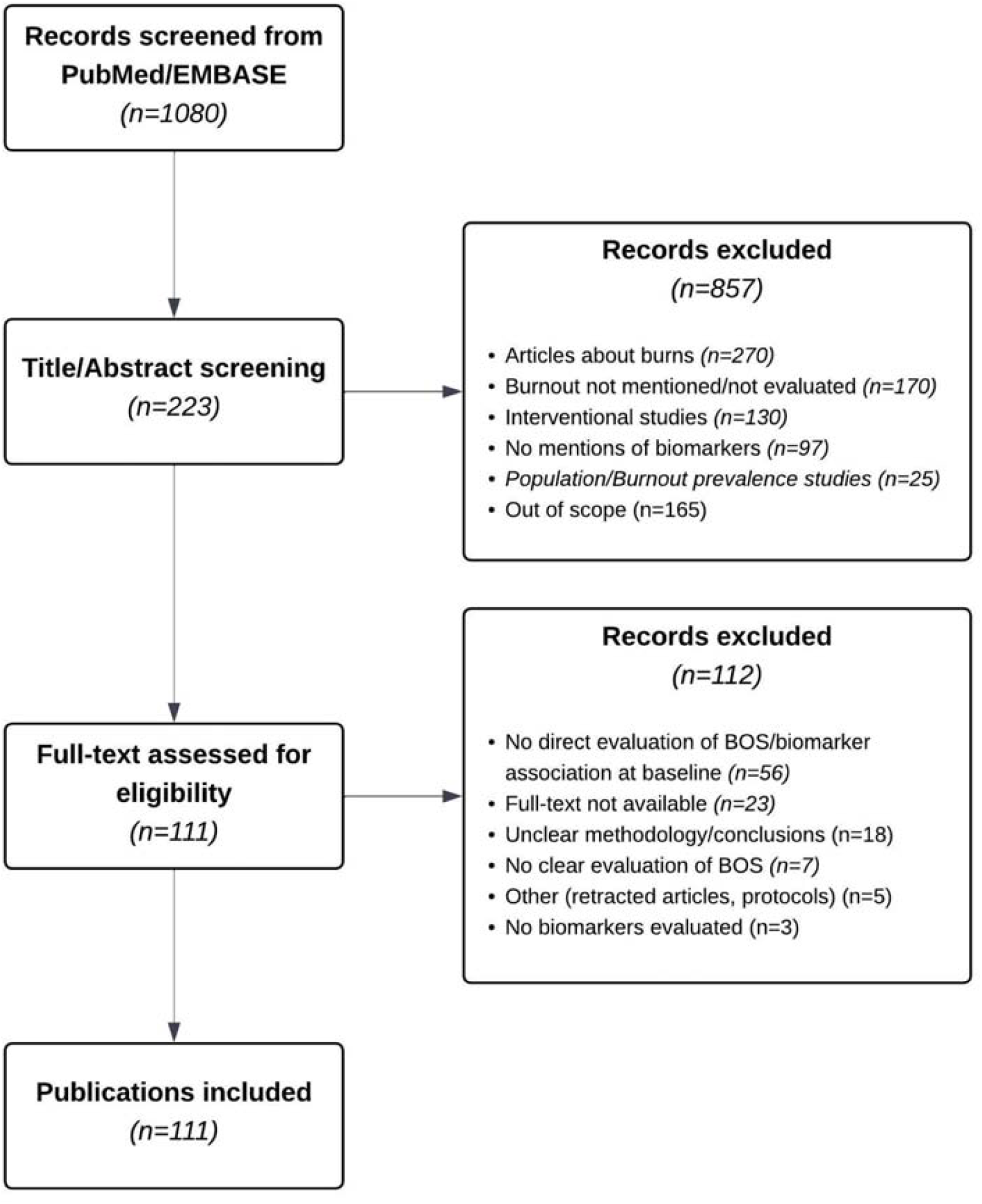
PRISMA Flowchart.

**Table 1.**
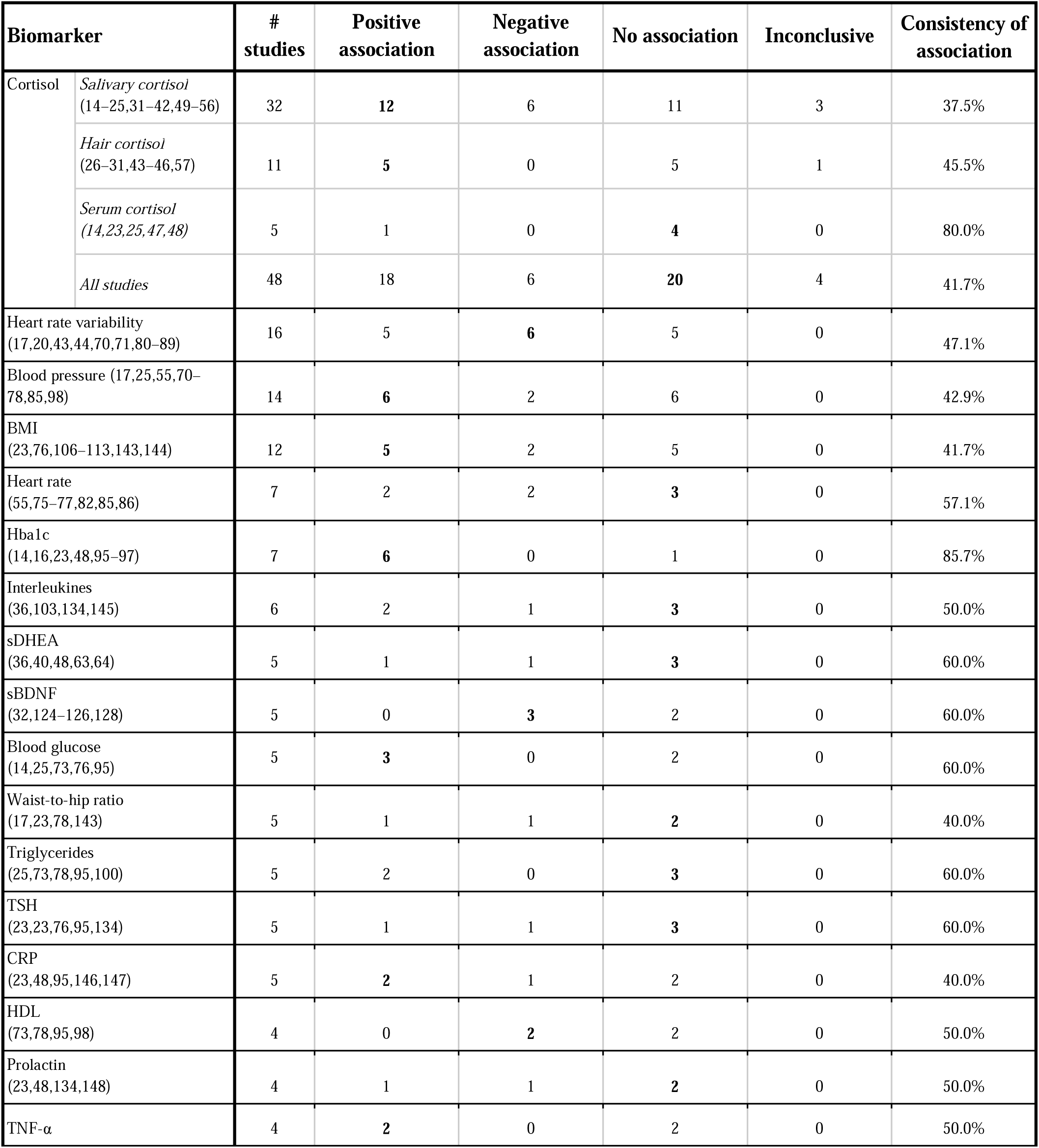
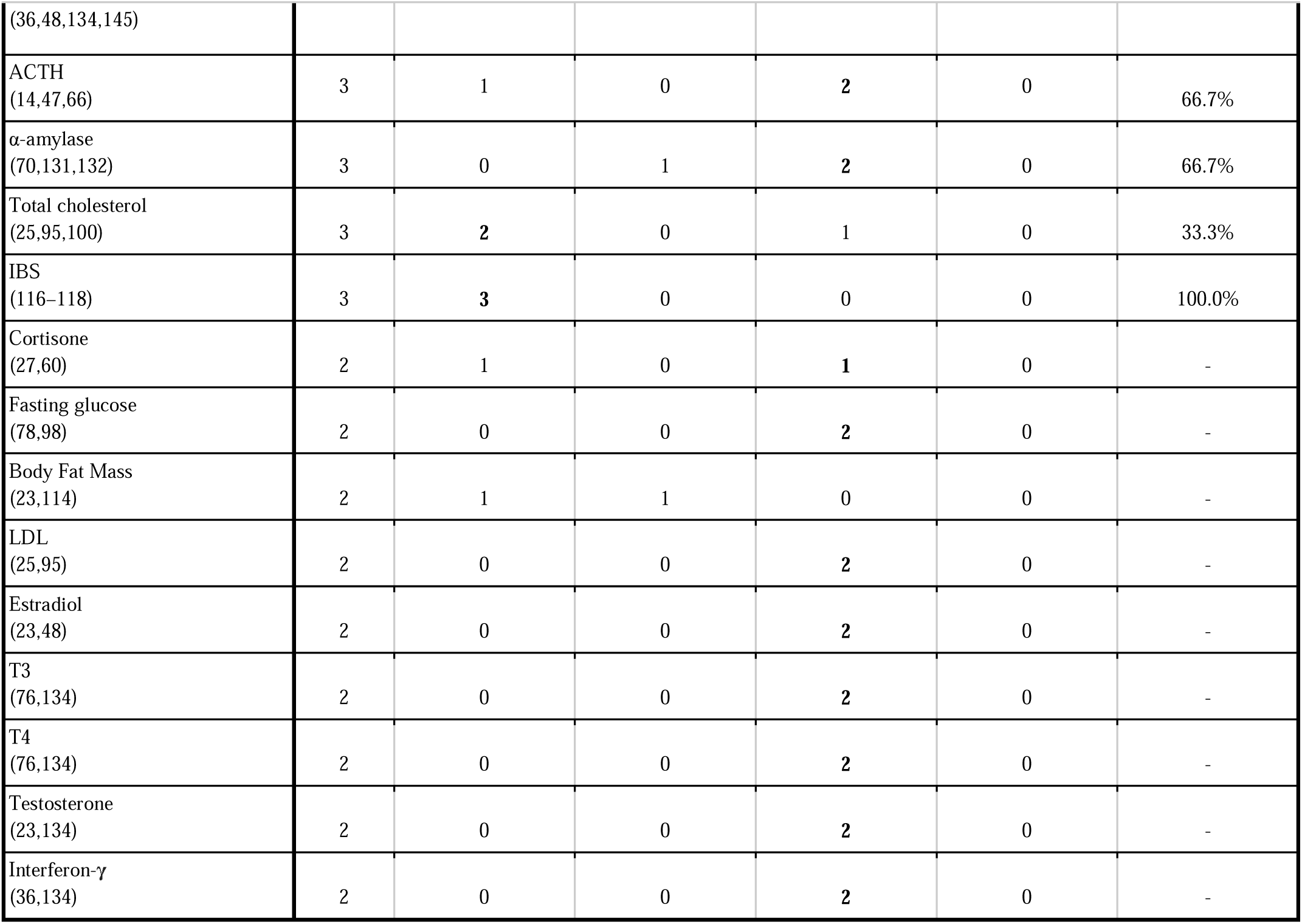
Frequency and consistency of the encountered biomarkers based on the systematic review of 111 articles.

| **Biomarker** |  | **# studies** | **Positive association** | **Negative association** | **No association** | **Inconclusive** | **Consistency of association** |
| --- | --- | --- | --- | --- | --- | --- | --- |
| Cortisol | *Salivary cortisol*<br>(14–25,31–42,49–56) | 32 | **12** | 6 | 11 | 3 | 37.5% |
|  | *Hair cortisol*<br>(26–31,43–46,57) | 11 | **5** | 0 | 5 | 1 | 45.5% |
|  | *Serum cortisol*<br>(14,23,25,47,48) | 5 | 1 | 0 | **4** | 0 | 80.0% |
|  | *All studies* | 48 | 18 | 6 | **20** | 4 | 41.7% |
| Heart rate variability<br>(17,20,43,44,70,71,80–89) |  | 16 | 5 | **6** | 5 | 0 | 47.1% |
| Blood pressure (17,25,55,70–78,85,98) |  | 14 | **6** | 2 | 6 | 0 | 42.9% |
| BMI<br>(23,76,106–113,143,144) |  | 12 | **5** | 2 | 5 | 0 | 41.7% |
| Heart rate<br>(55,75–77,82,85,86) |  | 7 | 2 | 2 | **3** | 0 | 57.1% |
| HbA1c<br>(14,16,23,48,95–97) |  | 7 | **6** | 0 | 1 | 0 | 85.7% |
| Interleukines<br>(36,103,134,145) |  | 6 | 2 | 1 | **3** | 0 | 50.0% |
| sDHEA<br>(36,40,48,63,64) |  | 5 | 1 | 1 | **3** | 0 | 60.0% |
| sBDNF<br>(32,124–126,128) |  | 5 | 0 | **3** | 2 | 0 | 60.0% |
| Blood glucose<br>(14,25,73,76,95) |  | 5 | **3** | 0 | 2 | 0 | 60.0% |
| Waist-to-hip ratio<br>(17,23,78,143) |  | 5 | 1 | 1 | **2** | 0 | 40.0% |
| Triglycerides<br>(25,73,78,95,100) |  | 5 | 2 | 0 | **3** | 0 | 60.0% |
| TSH<br>(23,23,76,95,134) |  | 5 | 1 | 1 | **3** | 0 | 60.0% |
| CRP<br>(23,48,95,146,147) |  | 5 | **2** | 1 | 2 | 0 | 40.0% |
| HDL<br>(73,78,95,98) |  | 4 | 0 | **2** | 2 | 0 | 50.0% |
| Prolactin<br>(23,48,134,148) |  | 4 | 1 | 1 | **2** | 0 | 50.0% |
| TNF-α | | 4 | **2** | 0 | 2 | 0 | 50.0% |

|  |  |  |  |  |  |  |
| --- | --- | --- | --- | --- | --- | --- |
| (36,48,134,145) |  |  |  |  |  |  |
| ACTH<br>(14,47,66) | 3 | 1 | 0 | 2 | 0 | 66.7% |
| α-amylase<br>(70,131,132) | 3 | 0 | 1 | 2 | 0 | 66.7% |
| Total cholesterol<br>(25,95,100) | 3 | 2 | 0 | 1 | 0 | 33.3% |
| IBS<br>(116–118) | 3 | 3 | 0 | 0 | 0 | 100.0% |
| Cortisone<br>(27,60) | 2 | 1 | 0 | 1 | 0 | - |
| Fasting glucose<br>(78,98) | 2 | 0 | 0 | 2 | 0 | - |
| Body Fat Mass<br>(23,114) | 2 | 1 | 1 | 0 | 0 | - |
| LDL<br>(25,95) | 2 | 0 | 0 | 2 | 0 | - |
| Estradiol<br>(23,48) | 2 | 0 | 0 | 2 | 0 | - |
| T3<br>(76,134) | 2 | 0 | 0 | 2 | 0 | - |
| T4<br>(76,134) | 2 | 0 | 0 | 2 | 0 | - |
| Testosterone<br>(23,134) | 2 | 0 | 0 | 2 | 0 | - |
| Interferon-γ<br>(36,134) | 2 | 0 | 0 | 2 | 0 | - |

**Table 2.** Frequency of the encountered BOS questionnaires based on the systematic review of 111 articles.

| Self-report questionnaire | Acronym | Number of occurrences with questionnaire | Proportions on studies using questionnaire |
| --- | --- | --- | --- |
| Maslach Burnout Inventory | MBI | 71 | 66.36% |
| Shirom–Melamed Burnout Questionnaire | SMBQ | 17 | 15.89% |
| Copenhagen Burnout Inventory | CBI | 4 | 3.74% |
| Oldenburg Burnout Inventory | OLBI | 4 | 3.74% |
| School Burnout Inventory | SBI | 3 | 2.80% |
| Diabetes Burnout Scale | DBS | 2 | 1.87% |
| Bergen Burnout Inventory | BBI | 2 | 1.87% |
| Stanford Professional Fulfillment Index | SPFI | 1 | 0.93% |
| Rome Burnout Inventory | RBI | 1 | 0.93% |
| Burnout Dimensions Inventory | BODI | 1 | 0.93% |
| Athlete Burnout Questionnaire | ABQ | 1 | 0.93% |
| Other questionnaires | - | 5 | 4.67% |

### Cortisol, sDHEA, ACTH

Cortisol, a glucocorticoid hormone, is considered a major and primary mediator of stress response (12,13). We identified 48 studies assessing the association between cortisol and BOS, exploring salivary cortisol, hair cortisol and plasma cortisol (respectively 66%, 23% and 11% of studies about cortisol). Amongst these articles, 18 studies reported a positive association (14,14–30). However, a substantial proportion of studies reported either no significant association (20 studies) (23,25,31–48), or even negative association (49–54) (6 studies) between cortisol and burnout. A few studies were considered inconclusive, as they described mixed associations across different BOS dimensions (31,55–57). When stratifying by cortisol sampling methods, similar proportions of association were found for salivary and hair cortisol, but not for plasmatic cortisol, which is more often found not to be associated with BOS (4 out of 5 articles). The large majority of articles (77.08%) used MBI to evaluate cortisol association to BOS. When the association was analyzed by BOS dimension (*i.e.* EE, DEP and PE) none showed a consistent association with a specific direction (Supplementary Table 3). Interestingly, 4 studies reported that patients with low BOS scores had higher salivary cortisol levels than those with high BOS scores (18,21,22,49). Two studies have also evaluated hair cortisone and the ratio between hair cortisol and hair cortisone, as cortisone might be a potential biomarker for free serum cortisol (58,59). One study concluded on a positive association (27) whilst the other found no association (60).

Whilst DHEA reflects the response to acute stressors, sulfate DHEA (sDHEA) has been shown to be linked with chronic stress (61). Accordingly, our review identified only studies investigating the association between sDHEA and BOS. Given its functional antagonism with cortisol, DHEA levels are expected to decrease under chronic stress conditions (62). However, 3 out of 5 studies did not show a significant association with BOS (40,48,63), whilst one showed a positive association (36) and the other a negative association (64). Interestingly, as DHEA is known to decrease with age, one study evaluated the correlation between sDHEA and BOS stratifying by age groups (64). A negative association was observed only in the 25-34 years old group, and was not observed in the whole BOS group vs. control group.

ACTH is the pituitary hormone of the HPA axis that stimulates the production of glucocorticoids, including cortisol and DHEA (65). We only included three studies on ACTH, of which two show no association to BOS (47,66), and one a positive association (14).

### Epinephrine, norepinephrine, aldosterone

Epinephrine and norepinephrine are key catecholamines released during the acute phase of the stress response, and are considered primary mediators of the AL (67,68). Despite their physiological relevance in stress adaptation, particularly in early or acute phases, no studies investigating their association with BOS were identified in our screening.

### Blood pressure, heart rate, and autonomic regulation

High blood pressure has been consistently used for the evaluation of AL, as chronic stress is a considerable risk factor for hypertension (67,69). 42.9% of screened articles found a positive association with systolic and/or systolic (SBP) and diastolic blood pressure (DBP) with BOS (17,70–74). An equal proportion of studies, however, found no correlation between blood pressure and BOS (25,55,75–78). Two studies reported statistically significant correlation with SBP and BOS but in opposite directions: a positive correlation in *De Vente et al.* (70) and a negative correlation in *Chico-Barba et al*. (78).

Heart rate variability (HRV), the variation of time interval between consecutive heartbeats, is considered to reflect the balance between parasympathetic and sympathetic nervous systems (79). Low HRV has been associated with high levels of stress, anxiety and depression (79). Among the 17 studies investigating HRV in BOS, 6 reported a negative correlation between HRV and BOS (70,80–84), consistent with a physiological distress and lack of resilience to psychological burdens (79). The other studies show either positive (17,20,71,85,86) or no correlation (43,44,87–89).

Heart rate (HR), also explored as a component of allostatic load, has been hypothesized to decrease with chronic stress exposure (90,91). However, most studies (3 out of 7) found no significant association between resting HR and BOS, suggesting that HR alone may lack sensitivity or specificity in this context (75,76,86).

### Glucose metabolism

Acute and chronic stress influences glucose metabolism, primarily through the activation of the HPA axis, leading to increased secretion of glucocorticoïds (GC) such as cortisol (92). Indeed, GCs promote hepatic gluconeogenesis and reduce peripheral glucose uptake, leading to transient hyperglycemia. Chronic exposure contributes to insulin resistance and increased risk of metabolic disorders (93). Elevated levels of glycated hemoglobin (Hba1c), blood glucose and fasting glucose have been associated with chronic stress (94). In our review, the majority of studies (85.7%) reported a positive correlation between Hba1c and BOS (14,16,48,95–97). The only study that found no correlation with Hba1c found however that the “reduced ability of distancing” dimension of Burnout Dimensions Inventory was associated with good insulin sensitivity (23).

Blood glucose tends to positively associate with BOS, as seen in 3 articles out of 5 exploring it (14,73,95). In contrast, fasting glucose and BOS were not correlated in the two studies concerned (78,98). These results indicate that glucose metabolism biomarkers are an interesting lead for BOS biomarkers.

### Lipid metabolism

Chronic high level of stress is associated with alterations in lipid profiles, including increased levels of total cholesterol (TC), triglycerides and decreased levels of high-density lipoprotein (HDL) (99). Indeed, both TC and HDL are commonly included in composite indices of AL. Regarding association with BOS, the screened studies show a similar trend: 2 out of 3 studies showed that total cholesterol tends to be associated positively with BOS (95,100) and 2 out of 4 studies showed that HDL negatively correlated to BOS (73,98), whilst the rest showed no correlation to BOS. No consistent association was observed for low-density lipoprotein (25,95), and triglycerides were mostly found to be unrelated to BOS (25,78,95). These results are based on few studies and would need further confirmation.

### Immunity and inflammation

Several markers of inflammation have been linked to acute and chronic stress (101). Higher interleukin-1b (IL-1b), interleukin-10 (IL-10), interleukin-6 (IL-6), stimulated interleukin-4 (IL-14) and interferon-γ (IFN-γ) are associated with acute and chronic stress (101). Indeed, IL-6 is sometimes used for AL (67,101,102). C-reactive protein (CRP) and Tumor Necrosis Factor-α (TNF-α) have been shown to be positively correlated with acute and chronic stress (101), however this association is challenged by another study (102). In our review, the association with inflammation markers (IL-1b, IL1-A, IL-2, IL-4, IL-6, IL-8, IL-10, IL-12, TNF-α, INF-γ) were rather non conclusive, with mostly no significant correlation with BOS. Interestingly, a study finds that associations of IL-12 and IL-6 with BOS are significant only in males (103).

### Obesity

Obesity is defined as a body mass index (BMI) over 30 kg/m^2^, has been shown to be intimately correlated to stress, as stress is a risk factor for obesity (104), with cortisol as a mediator (105). We observe a trend in this direction, with 5 out of 12 studies showing a positive correlation of BMI with BOS (106–110). However, there are also 5 studies showing no correlation to BOS (23,76,111–113). Another corpulency measure, waist-to-hip ratio (WHR), has been associated with stress (104), and is included in the AL measure (67,68). Other corpulency measures might benefit from more investigation; body fat mass data was inconclusive (23,114), whilst lean body mass was found to be positively correlated to BOS in one study (23).

### Irritable bowel syndrome

Irritable bowel syndrome (IBS) is a chronic or fluctuating dysfunction of the gastrointestinal tract, which roots in diverse origins such as immune, hormonal and nervous systems malfunctions (115). Acute and chronic stress has been shown to have an impact on intestinal sensitivity and be a crucial factor in the development of irritable bowel syndrome (IBS) (115). We observed that the three articles screened on IBS show a positive association with BOS (116–118).

### Allostatic load

We found two articles assessing the correlation between AL and BOS (119,120). They differed in their conclusion, *Juster et al.* found a positive correlation (119), and *Langelaan et al*. found no correlation (120). However, both studies did not use the same list of biomarkers for their AL measure, which can explain this discrepancy. Whilst *Langelaan et al*. focused on a shortlist of biomarkers, close to the original AL list (SBP, DBP, BMI, CRP, total cholesterol, HDL, Hba1c), *Juster et al.* explored a more extensive list of biomarkers (Salivary cortisol, DHEA, CRP, fibrinogen, insulin, SBP, DBP, Hba1c, albumin, creatinine, pancreatic amylase, total cholesterol, HDL, triglycerides). This boils down to the fact that many variations of the AL biomarkers list have been used and there is yet to reach a consensus in terms of biomarker selection (121).

### Brain-derived neurotrophic factor (BDNF)

Brain-derived neurotrophic factor (BDNF) is a neurotrophin involved in neuronal survival, synaptic plasticity, and stress resilience (122). High chronic stress was potentially linked to low basal serum BDNF (sBDNF) levels (123). We observed that in 3 studies, sBDNF was negatively associated with BOS (32,124,125) whilst 2 other studies show no correlation to BOS. Other aspects of BDNF, *i.e.* promoter methylation and gene polymorphism have also been evaluated with regard to BOS association (126–128). Different gene polymorphism can have different associations to BOS. *Jia et al.* (127) showed that rs6265 had a positive correlation to BOS whilst *He et al.* (128) showed no correlation of rs2049046 to BOS. BDNF promoter methylation, which is expected to downregulate sBDNF levels, was found to be positively associated with BOS (126). In spite of this, BOS was not associated with alterations of sBDNF levels in this study. This points out to the necessity to investigate these mechanisms more in depth.

### Other hormonal biomarkers

Beyond its role as a lactation hormone, some evidence points to acute and potentially chronic stress association to prolactin (129). However, associations found in our study do not allow us to conclude for its association to BOS.

Salivary alpha-amylase (sAA) has been proposed recently as a strong candidate biomarker for acute and chronic stress (130). In the 3 studies concerning sAA in our review, 2 showed no correlation to BOS (131,132) and one showed a negative correlation to BOS (70).

Thyroid-stimulating hormone (TSH) has been shown to tend to correlate positively with stress, although not significantly (133). Interestingly, in *Kautzky et al.*, TSH was found to be associated with BOS, but the direction of the association depended on gender. Two Burnout Dimension Inventory dimensions (“reduced ability of distancing” and “dysfunctional compensation”) were positively correlated to BOS in females, whilst a negative correlation to BOS was observed in males (23). However, in the other articles selected in our research, we didn’t observe a consistent association of TSH to BOS (76,95,134). We did not observe correlation with BOS of estradiol and testosterone, nor T3 or T4 (23,48,76,134).

## Discussion

In this systematic review, the association of BOS with 36 potential biomarkers has been evaluated. Only Hba1c, blood glucose and IBS showed relatively consistent patterns of association with BOS. In contrast, many other physiological indicators, including cortisol, inflammatory cytokines, and lipid markers, demonstrated inconsistent or inconclusive findings. This variability likely reflects both biological heterogeneity within BOS and methodological discrepancies across studies.

Even though the concept of burnout exists for a few decades, BOS definition remains an object of controversy. Most studies use the MBI to define the BOS status of a patient, whilst the MBI is disputed in the literature as narrowly framing burnout into an inconsistent syndrome (4,5). The developers of the MBI indicate that the questionnaire shouldn’t be used to calculate based on the sum score of its three dimensions (135). We observed that in studies that evaluate EE, DEP and PE separately, the direction of the associations with the evaluated biomarkers are not always coherent among the three dimensions, leading to results that are difficult to integrate and interpret (29,32,49,78,108,116,136). PE typically exhibits associations that diverge from those observed with EE and DEP (Supplementary Table 3). We also observed that for a number of studies, EE was the sole dimension used to assess BOS, as considered by many the most important aspect of BOS (Supplementary Table 3). When compared to the whole BOS score, EE’s association was coherent with the whole BOS score association (14,14,14,14,26,81). As EE correlates highly with depression (130), and is in fact a basic symptom of depressive conditions (under such labels as loss of energy, fatigue, and dysphoric mood), *Bianchi et al.* have suggested that job related-distress should be approached through the prism of occupational depression rather than that of burnout (138). Another avenue to be explored could be to consolidate BOS evaluation with the addition of cognitive impairment, since it was integrated to BOS conceptualisation (7–9). Ultimately, redefining BOS definition and measurements might be able to pinpoint more coherently eventual BOS correlation with biomarkers.

In our review, we observed that cortisol, as well as other primary mediators and effectors of the stress response, are not clearly associated with BOS. One of the potential mitigating factors could be the adaptation of the cortisol response to long term stress, and therefore burnout status. Hypercortisolism observed in stress responses can, over a long period, change into hypocortisolism, as a possible preservation mechanism of the HPA axis (65). This could explain why we observe higher levels of cortisol in low BOS patients compared to high BOS patients. This has also been observed regarding the CAR responses (20,49–51,53,56), for which CAR response was found flattened in high BOS patients compared to controls or low BOS, corresponding to a maladaptive response from the HPA axis (139). These results could indicate that cortisol levels could only accurately be used as a marker for early stages of BOS. Another aspect of this question is raised by *Traunmüller et al.* (17) who identify in the BOS patients two subpopulations, “illusory” BOS and “coherent” BOS. The “illusory” BOS subgroup has a less consistent association with biomarkers than the “coherent” group, as well as lower BOS scores and higher association with depression. Knowing that depression has been associated with hypocortisolism (140–142), this could be another explanation for different levels of cortisol in the BOS population, highlighting the need to better understand its heterogeneity. This classification of BOS population reverberates to other biomarkers, whose alterations are directly related to cortisol and the HPA axis.

Another angle to tackle the lack of correlation between biomarkers and BOS is how the association is assessed. Most studies will look at correlations or simple/multivariate linear regression, whilst other types of association between these variables could be explored. An example would be *Wendsche et al*., in which no correlation was found between BOS and salivary cortisol, however the exponential term of MBI positively correlated with BOS (57). This highlights the importance of investigating non-linear relationships between biomarkers and BOS.

This systematic review highlights a persistent difficulty in identifying the biological underpinnings of BOS. While certain candidate biomarkers, such as HbA1c, blood glucose, and irritable bowel syndrome, appear to show more consistent associations with BOS, no clear biological pattern has yet emerged. Most physiological systems investigated, including the HPA axis, inflammatory pathways, lipid metabolism, and neuroplasticity markers (BDNF), yielded inconclusive or contradictory findings.

These inconsistencies may partly reflect methodological variations, but they also point to a deeper conceptual issue: the lack of a unified, clinically grounded definition of BOS. The overreliance on the MBI, with its psychometric limitations, likely contributes to the observed discrepancies across studies. Alternative measures, such as the Occupational Depression Inventory, may offer a more valid and clinically meaningful framework for capturing work-related psychological distress.

From a clinical perspective, our findings suggest that current biological evidence does not yet support the routine use of biomarkers for BOS diagnosis or staging. However, early alterations in metabolic, immune, or neuroendocrine markers may help identify vulnerable individuals, particularly in occupational health settings. In this perspective, improving the early identification of burnout is essential not only for advancing clinical research, but also for informing timely and targeted preventive strategies.

## Data Availability

All information are available in supplemental files.

## Acknowledgments

We are grateful to all co-authors for their complementary expertise and to our clinical and data collaborators for their valuable input throughout the project.

## Disclosures

Financial interests: F.Z. and M. B were compensated for their assistance in manuscript preparation and medical data review by Zoī SAS. Non-financial interests: the authors declare no other relevant non-financial interests to disclose.

## Notes

### Funding Statement

This study did not receive any funding

